# Modeling the systemic risks of COVID-19 on the wildland firefighting workforce

**DOI:** 10.1101/2021.09.15.21263647

**Authors:** Erin J Belval, Jude Bayham, Matthew P Thompson, Jacob Dilliott, Andrea G. Buchwald

## Abstract

Wildfire management in the US relies on a complex nationwide network of shared resources that are allocated based on regional need. While this network bolsters firefighting capacity, it may also provide pathways for COVID-19 transmission between fire sites. We develop an agent-based model of COVID-19 built on historical wildland fire assignments using detailed dispatch data from 2016-2018, which form a network of firefighters dispersed spatially and temporally across the US. We use this model to simulate SARS-CoV-2 transmission under several intervention scenarios including vaccination and social distancing. We find vaccination and social distancing are effective at reducing transmission at fire incidents. Under a scenario assuming High Compliance with recommended mitigations (including vaccination), infection rates, number of outbreaks, and worker days missed are effectively negligible. Under a contrasting Low Compliance scenario, it is possible for cascading outbreaks to emerge leading to relatively high numbers of worker days missed. The current set of interventions in place successfully mitigate the risk of cascading infections between fires, and off-assignment infection may be the dominant infection concern in the 2021 season. COVID-19 control measures in place in wildfire management are highly beneficial at decreasing both the health and resource impacts of the ongoing pandemic.

## Introduction

The wildland firefighting system in the United States (US) saw unprecedented challenges in 2020 as the COVID-19 pandemic added additional complexity to a severe fire season^1^. Concerns about COVID-19 outbreaks at individual fires^2,3^ spurred the development of COVID-19 prevention and mitigation procedures including how fire camps were operated and how firefighters interacted with each other^4^. The implications of a COVID-19 outbreak on a single fire have been modeled^3^, but potential system-wide impacts have not yet been explored^5^. Here we explore potential health and workforce capacity impacts by modeling the movement of wildfire suppression resources across the country over an entire fire season and the corresponding potential for disease spread and cascading outbreaks across wildfire incidents.

While COVID-19 vaccination will mitigate transmission of COVID-19 in the 2021 fire season, there is uncertainty surrounding the level of personnel that might be vaccinated^6^ and, particularly early in the season, COVID-19 impacts to the wildland firefighting workforce are still of concern^7^. There is a compelling reason to explore these potential impacts; degradation of workforce capacity and operational readiness were acutely felt at times during the 2020 fire year. For example, the Cameron Peak fire saw 76 SARS-CoV-2 (the virus that causes COVID-19) infections and more than 250 personnel isolated over the course of the incident, which saw days with over 1000 personnel assigned to the fire^8^.

Wildland firefighters, particularly those working on large fires, are a highly transient workforce. Regions with low or moderate fire activity allow some of their firefighters to be reassigned to other regions that need additional firefighting capacity^9^. For example, firefighters from the Southwestern region are often used to support fires in the Northern Rockies because the peak fire seasons differ across the regions. Figure 1 depicts the incoming assignments originating all over the country and outbound reassignments to a particular fire in Montana. These cross-boundary assignments provide flexibility in wildfire response capacity as single incidents can require thousands of personnel, however they also pose a potential threat in the context of infectious disease spread. Reassignments from one fire to another often happen within a few days, thus, an outbreak of disease at one fire has the potential to spread to other fires. These cascading effects can accelerate SARS-CoV-2 spread across the national wildland firefighting workforce as the fire season progresses. In addition to the health risks associated with SARS-CoV-2 outbreaks, multiple fires with outbreaks could lead to resource deficits, with a sizable portion of firefighters out sick or quarantined^8^. Because the firefighting workforce is finite and, at the height of the fire season, some requests for firefighters go unfilled^10^, losing a portion of the workforce to sickness and quarantine is a significant concern. Therefore, there is a need for model-based assessment of COVID-19 risk at the national, seasonal scale.

**Figure 1:**
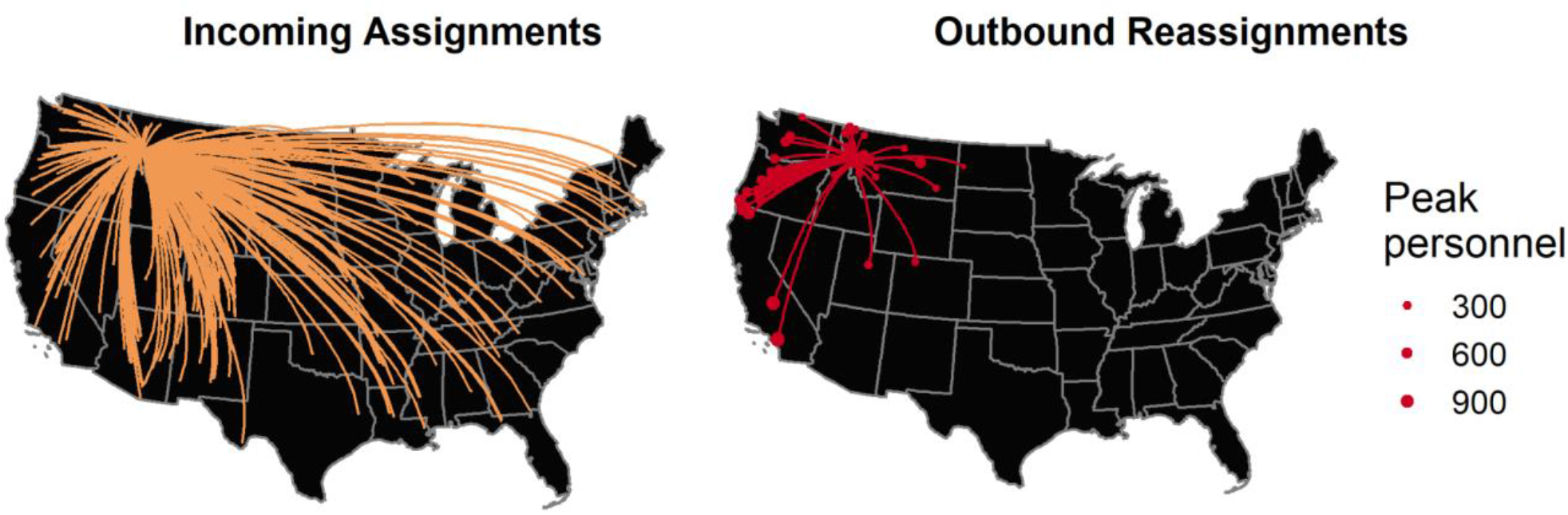
Historical assignment/reassignment data for a single fire in Montana. The map of incoming assignments shows the range of origins for personnel assigned to a fire that started on July 15, 2017. The outbound reassignments shown include all incidents to which personnel went, given nine or fewer days between demobilization at the first fire and mobilization at the second fire.

Agent-based models (ABMs) have been adapted to model the spread of SARS-CoV-2 for a variety of settings. ABMs have been used to describe SARS-CoV-2 spread within cities^11,12^ and at the national level^13,14^ primarily to describe disease dynamics and examine the potential impact of various intervention strategies^15^. They have also been used to identify locations at high risk of driving infection outbreaks, and to simulate SARS-CoV-2 spread between locations^16^. An ABM is the ideal tool to examine infection spread within the wildland fire response community as it allows for explicit modeling of interactions between individuals and can track the movement of individuals between fire locations.

We develop an epidemiological ABM to simulate the transmission of SARS-CoV-2 across the wildfire response system based on actual historical assignment data to study the potential impacts of the pandemic on wildfire response capacity throughout the season. We use the model to simulate several mitigation measures including the so-called “module-as-one” (i.e., pods) policy, social distancing, and vaccination. Figure 2 describes the mechanics of the model and the interaction between firefighters assigned to an incident. Crew modules consist of crew personnel who have high levels of contact within their module but are largely isolated from other modules. To simulate this, in our model each crew has a set of personnel who are designated leaders that interact with management personnel and other module leaders. If any individual in a module is diagnosed with COVID-19, then the entire module quarantines^5^, as all module members are assumed to be in high levels of contact with each other. Management personnel are unable to act as a self-contained module because they need to interact with many management and crew personnel to coordinate wildfire operations^5^. Management personnel isolate when diagnosed with COVID-19, but there is no quarantine of others associated with the diagnosis in our model. SARS-CoV-2 can spread within a fire as described in Figure 2 and between fires as those firefighters are reassigned to other fires across the country throughout the season (Figure 1). Firefighters may also contract the disease while off-duty based on the rate of community transmission. We provide more details on the model in the Methods section and the supplementary materials. The granularity of the model allows us to investigate the burden of COVID-19 as well as its impact on workforce capacity on multiple scales from individual fires to the system as a whole.

**Figure 2.**
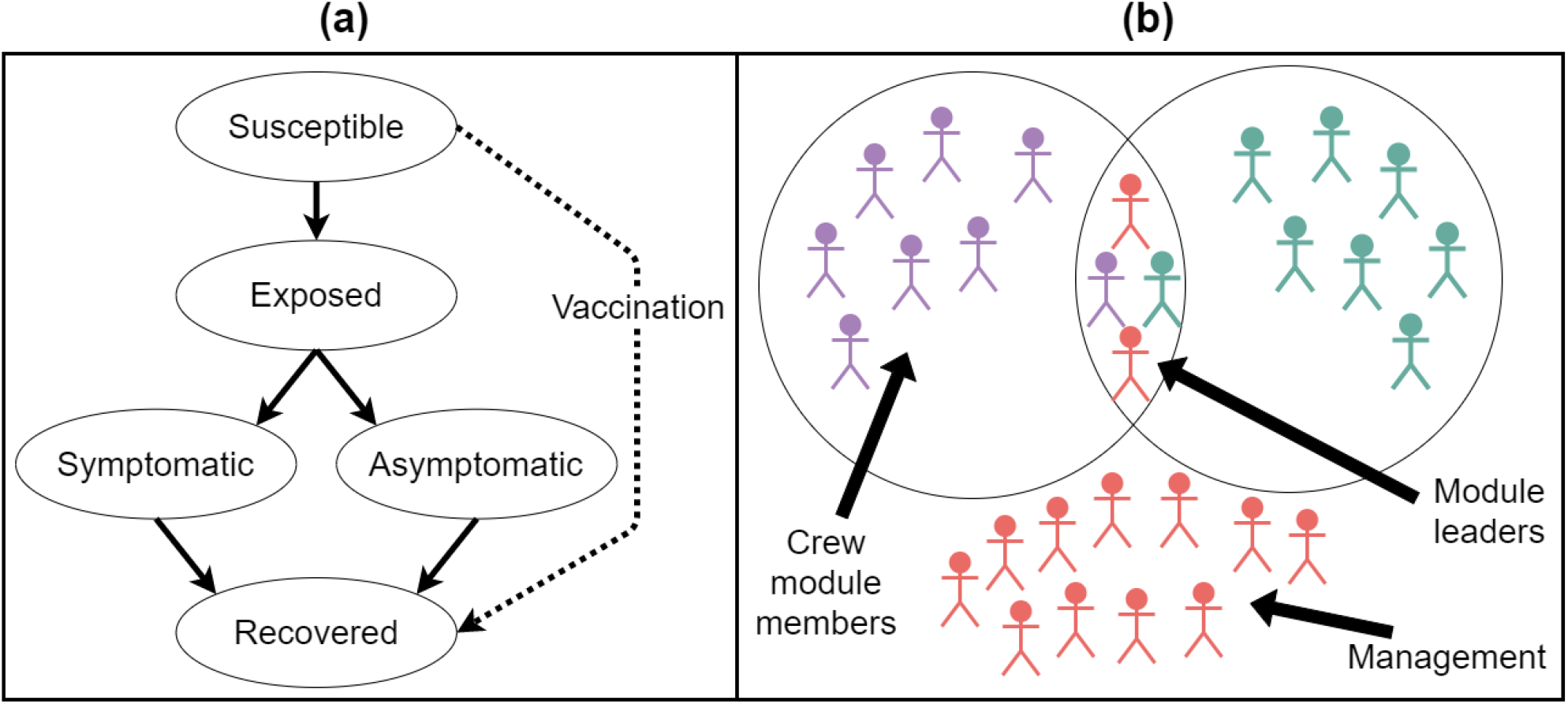
(a) The possible viral states which individuals may travel through in simulations. The arrows indicate possible paths that individuals may take through the viral states. An individual may move directly from susceptible to recovered only if vaccinated. (b) Interactions between personnel on a single fire. Crew module members (individuals of the same color) interact only with other members of the same module, with the exception of module leaders, who interact both with their module members and with other module leaders. Management personnel cannot effectively form modules and thus interact with all other management personnel as well as a proportion who interact with the crew module leaders.

## Results

We develop scenarios to address two key uncertainties in the interplay between the fire season and the COVID-19 pandemic: vaccination and social distancing behaviors of wildland fire personnel and the spatio-temporal variation of fire occurrence. We address the uncertainty in vaccination rate and compliance to social distancing behaviors among wildland fire personnel by creating three distinct behavioral scenarios: a low behavior compliance scenario, a baseline scenario, and a high behavior compliance scenario. The “Low Compliance” scenario assumes less compliance with infection control measures (i.e., low effort to maintain social distancing and lower percentages of individuals correctly diagnosing their symptoms) and fewer vaccinated individuals. The “High Compliance” scenario assumes more compliance with social distancing, more frequent diagnosis of symptoms, and more vaccinated individuals. The “Baseline” scenario assumes a moderate level of social distancing compliance, symptom identification, and vaccination. The specific parameters used for each scenario can be found in the supplementary materials. We address the variation in fire occurrence patterns by using fire assignments from three distinct fire seasons: 2016, 2017, and 2018. These years cover a range of spatial and temporal demand for wildland fire suppression resources.

We simulate the model 100 times in each scenario (Baseline, High Compliance, Low Compliance) for each fire season to illustrate the uncertainty due to stochastic transmission and yearly variation in firefighter assignments. We focus on four outcomes relevant to the wildfire management community: 1) the number of cumulative infections over the season, 2) outbreaks of COVID-19 on individual fires, 3) reassignments of infectious personnel between fires, and 4) workforce absenteeism due to quarantine. We report median values of the 100 simulations along with the interquartile range (IQR; indicates the central 50% of the distribution).

Figure 3 illustrates the number of cumulative infections contracted both on and off of active duty across the three scenarios over the duration of the season using 2017 fire assignment data. There were 43,360 personnel assigned to at least one large fire in 2017. Figure 3 shows that the number of infections acquired off-fire is substantially more than those acquired on-fire. Using only the Baseline results across all three years, our results suggest that approximately 95% of infections are acquired while firefighters are off duty in these scenarios. Many of these infections acquired off duty may be asymptomatic and pose a risk to other firefighters once the infected individual is deployed to an incident. The Low Compliance median percentage of personnel acquiring SARS-CoV-2 off-fire is about 9.4%. Figure 3 also shows that transmission on fires closely follows wildfire activity over the course of the season. In the Low Compliance scenario, cumulative infections on fire rise rapidly during the summer (days 150-250) and plateau once fire activity decreases. In Figure 3 we can observe that the High Compliance scenario effectively reduces transmission relative to the Baseline and, similarly, the Baseline scenario reduces transmission relative to the Low Compliance scenario. Specifically, the median number of cumulative infections for the Baseline scenario for 2017 is 1915 [IQR: 1892-1944] while the median of the High and Low Compliance scenarios are 634.5 [IQR: 618.8-654.2] and 4512 [IQR: 4461-4566], respectively.

**Figure 3:**
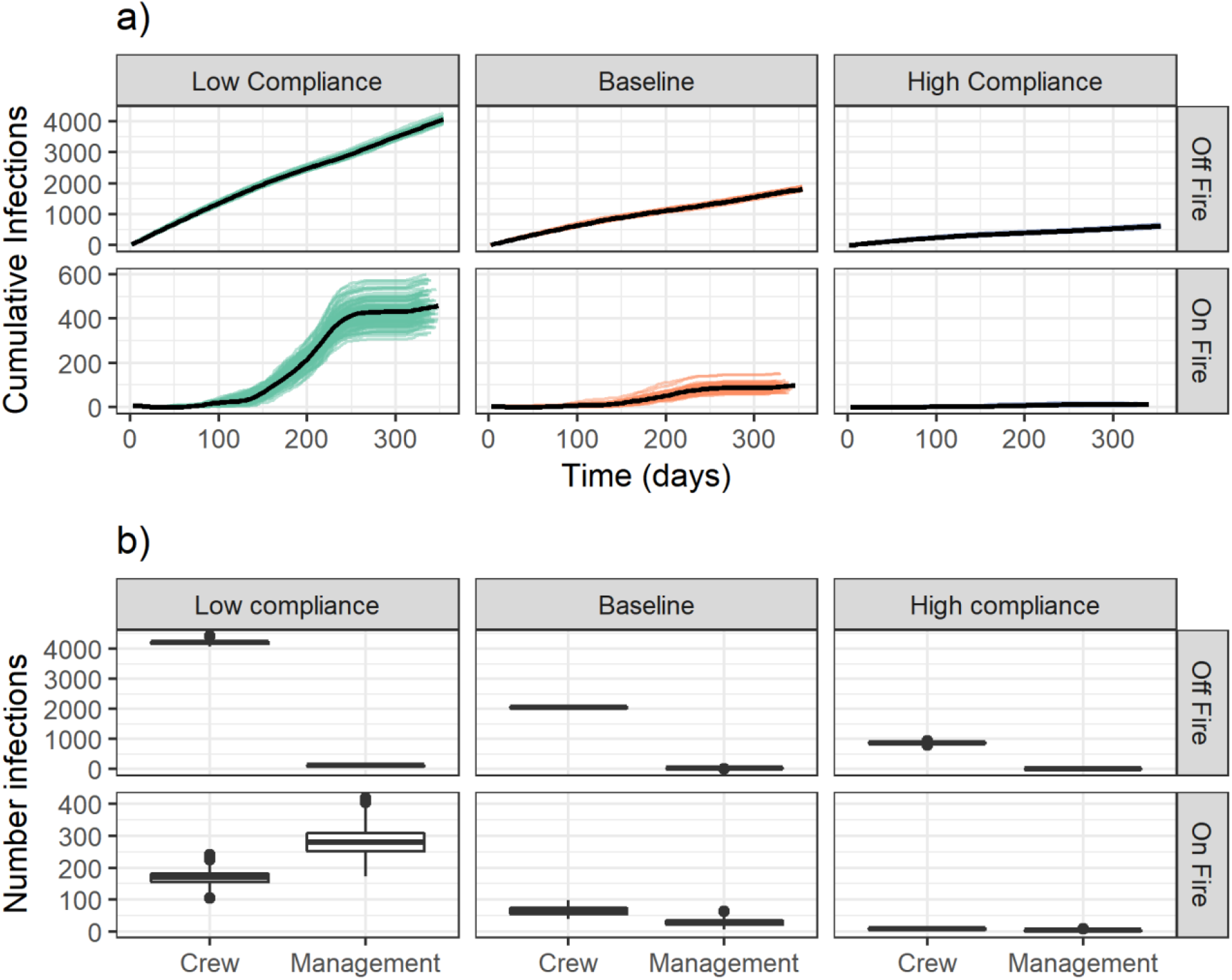
Daily cumulative infections by compliance scenario on and off fire (a) and annual cumulative infections by personnel type (b). In (a), each line is associated with a single scenario run while the bolded lines show the run with the median number of cumulative infections incurred. The total cumulative infections across the 2017 season by scenario and personnel role are shown in (b), with cases attributed to assignment status at time of exposure.

The number of contacts and the intensity of those contacts is not homogeneous across personnel. This is reflected in our model structure and parameters (see the Materials section and supplementary materials for details). We therefore examined the number of on-fire infections that occurred specifically within management personnel and crew personnel modules (Figure 3b).We find that in the Low Compliance scenario, there is a relatively high ratio of management to crew infections for cases incurred on a fire as compared to those incurred off fire (a median of 279.5 (IQR: 251.8-308.8) and 169.5 (IQR: 155.8-182.2) infections incurred on fire and a median of 121 (IQR: 112-128) and 4205 (IQR: 4161-4234) infections incurred off fire by management and crew, respectively). As compliance with mitigation measures increases, the ratio of management to crew on-fire cases goes down. This likely reflects the contact structure for management personnel (they are exposed to more people each day) and the isolation procedures (only the symptomatic person isolates if they are management as opposed to the entire module for crew personnel). The implications of the higher caseloads incurred on fire by management personnel has significant implications. First, management personnel tend to be older than crew personnel, which means they are also at higher risk of severe symptoms. Second, key management positions require high levels of qualifications, so higher caseloads in management personnel may burden the wildland firefighting system more than caseloads in crew personnel.

The spatio-temporal variation in fire activity between seasons did not substantially affect simulated cases of SARS-CoV-2 incurred on fire across the Baseline scenario. The median number of cumulative infections for runs using the assignments from the years 2016, 2017, and 2018 under the Baseline scenario assumptions was 79.5 [IQR: 72-88], 94 [IQR: 81-102], and 94 [IQR: 82.75-108.25] respectively. We do observe a slightly higher level of cumulative simulated infections overall using 2017 and 2018 assignments than those from 2016; this is because the total number of personnel assigned to a large fire was higher in the 2017 and 2018 scenarios, leading to a larger pool of personnel that can be infected off fire. The median number of cumulative infections using 2016 assignments was 1498 [IQR: 1471-1521], using 2017 assignments was 1915 [IQR: 1892-1944], and using 2018 assignments was 1808 [IQR: 1782-1849]. Further exploration of the differences between scenarios can be found in the supplementary materials. Because there was little variation in disease spread patterns by assignment-year in on-fire infections, we focus the rest of our results on scenarios based upon the 2017 fire assignments.

While the number of individual cases are an important systemic outcome, outbreaks of COVID-19 on a wildfire incident can add substantial burden on the management team. Therefore, for each run we counted the number of cases of SARS-CoV-2 on each fire. If a fire incurred at least two cases from different crew modules, two management personnel with cases, or a combination of crew and management personnel with cases, we counted that fire as having an outbreak for that run. Figure 4a shows the percentage of runs for which each incident had an outbreak by the maximum number of personnel assigned to the fire on a single day. We find that the incidents most likely to see outbreaks are the incidents with the highest number of maximum personnel assigned. Compliance with interventions has a greater impact the larger the number of personnel on the fire. While the maximum number of personnel on the fire has a strong relationship with the percentage of runs in which each fire experiences outbreaks, duration of the fire also plays an important role. We single out two fires in Figure 4a: the points associated with one fire are circled in blue (the “many-outbreaks fire”) and the points associated with the second fire have pink squares around them (the “fewer-outbreaks fire”). When we examine the number of personnel on the fire over time (Figure 4b), we see that the many-outbreaks fire lasted much longer than the fewer-outbreaks fire.

**Figure 4:**
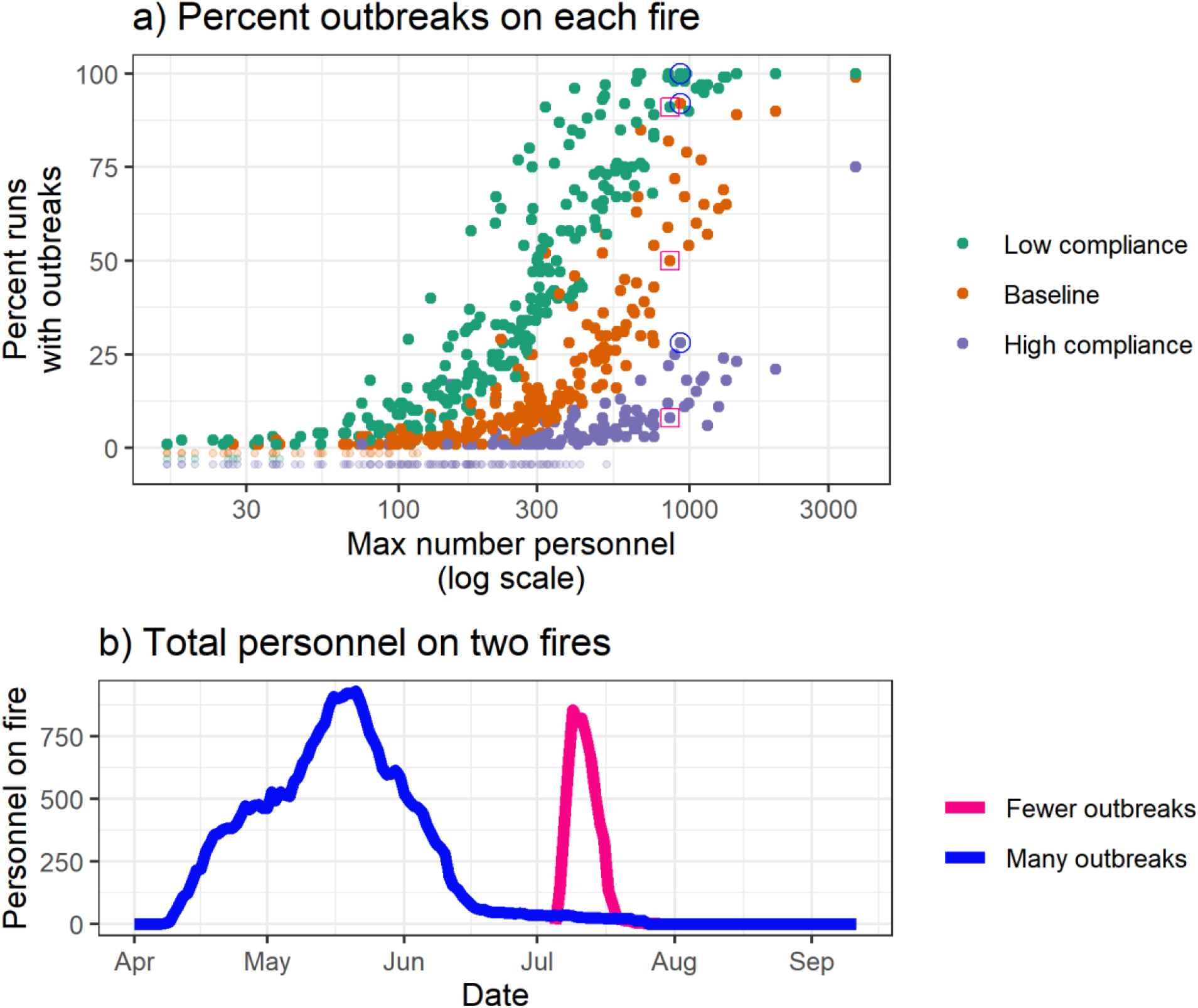
a) Percentage of runs for which each fire had an outbreak by scenario and maximum number of personnel assigned to the fire on a single day. Two fires are singled out: the points associated with a “many outbreaks” fire are circled in blue and the points associated with a “fewer outbreaks” fire have a pink square around them. b) The number of personnel over time for the “many outbreaks fire” and the “fewer outbreaks fire” that are indicated in (a).

To explore the risk of personnel transmitting disease from one fire to another, we examined the number of infectious assignments and reassignments. These metrics provide a way to quantify the difference in risk from personnel contracting the virus off fire and bringing it to their assignment versus the risk from personnel bringing the virus from one fire to another. We find that the number of infectious assignments from personnel who contracted SARS-CoV-2 off fire is higher than the number of infectious reassignments from personnel who went from one fire to another while in an exposed or infectious state (Figure 5a). Management personnel have a relatively high risk of being reassigned while infectious relative to the number of infectious assignments they have, particularly in the Low Compliance scenario (121 [IQR: 112-128] and 161 [IQR: 154.8-173] infectious assignments and 28 [IQR: 17-47] and 33 [IQR: 26.75-41.25] infectious reassignments for management and crew, respectively, in the Low Compliance scenario).

**Figure 5.**
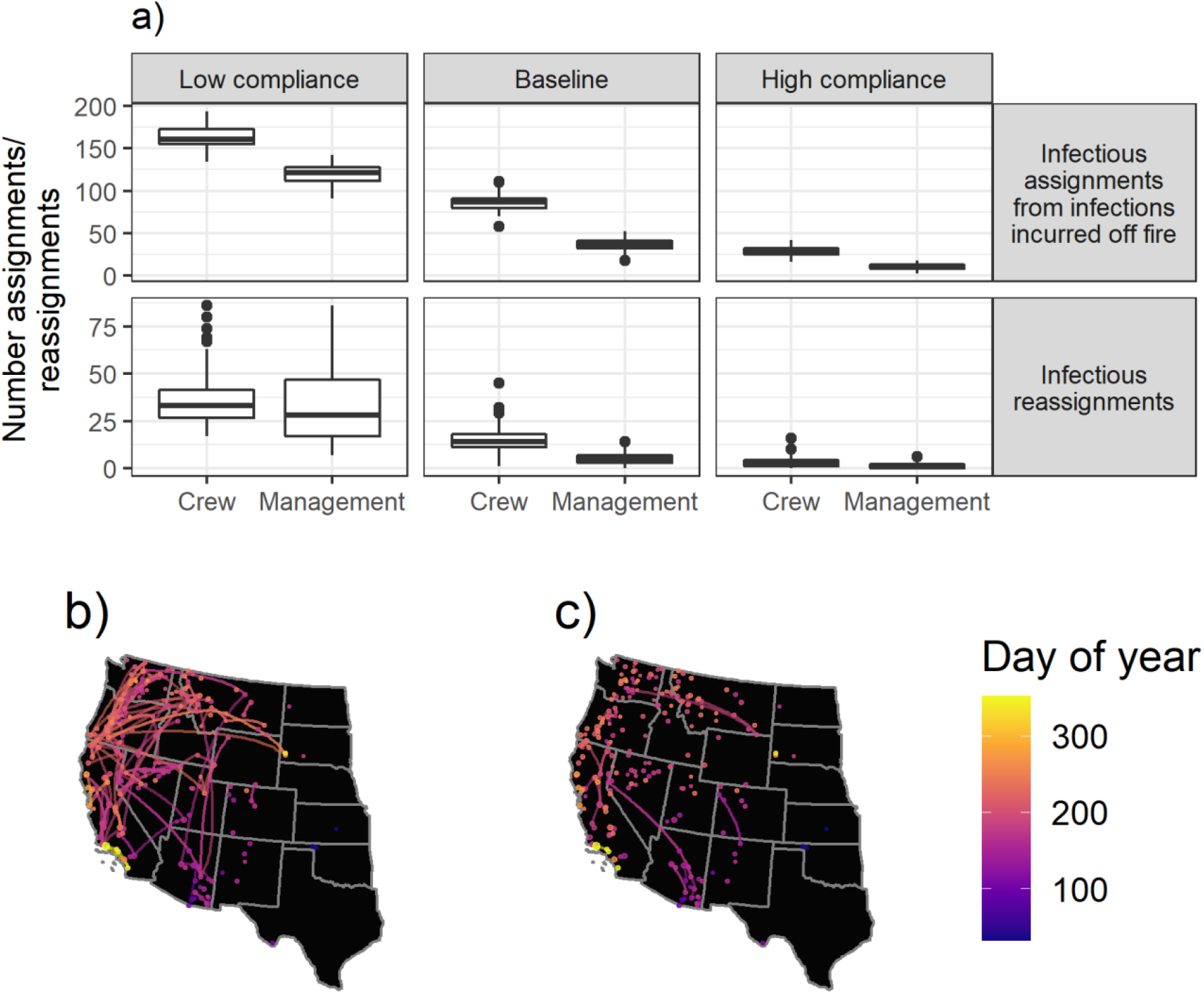
a) The number of infectious assignments and reassignments by scenario and personnel type for the 2017 fire assignment data. b) A map of the infectious reassignments that occurred during the Low Compliance run using 2017 data that had the highest number of infectious reassignments (i.e., the worst case scenario observed). c) A map of the infectious reassignments that occurred during the High Compliance run using 2017 data that had the highest number of infectious reassignments. All large fires included in the analysis are mapped as points, with the point size corresponding to the maximum number of personnel assigned to the fire on a single day. Lines connecting fires indicate infectious reassignments.

A comparison of two specific runs illustrates the effectiveness of mitigation measures in reducing infectious reassignments. A map of the worst case scenario for reassignments (i.e., the highest number of infectious reassignments observed) in the Low Compliance scenario is shown in Figure 5b, while a map of the worst case scenario for infectious reassignments in the High Compliance scenario is shown in Figure 5c. In the Low Compliance worst case scenario, we can observe disease being transferred between fires across space and time, while in the High Compliance worst case scenario we see many fewer infectious reassignments.

In addition to the health of firefighting personnel, agency administrators are concerned with workforce capacity and the ability to accomplish firefighting objectives. When a firefighter self-identifies as infected, that individual’s module is quarantined to reduce transmission. However, vaccinated individuals are not required to quarantine after exposure under current guidance^5^. Figure 6 compares the number of firefighter days missed by scenario, showing the number of days that individuals that would be required to quarantine given no vaccination (that is, all individuals quarantine regardless of vaccination state) and the number of days that individuals that are actually required to quarantine (i.e., vaccinated individuals are excluded). In the Baseline scenario, SARS-CoV-2 exposure and quarantine leads to 1007 [IQR 842-1198] firefighter days missed, which represents less than 1% of total assigned days (1918 [IQR 1718-2354] if vaccinated individuals are required to quarantine). As a point of comparison, the Cameron Peak Fire alone could have accounted for more than 2,000 worker days missed^8^. The median number of worker days missed for the Baseline scenario is slightly lower than the median of the Low Compliance scenario (1346 [IQR: 1081-1572]). The High Compliance scenario yields the fewest worker days missed (240.5 [IQR: 187.8-310.8]), but the distribution shows that higher impacts on workforce capacity are possible, highlighting the uncertainty faced by fire managers throughout the pandemic. We summarize worker days missed in each of the mitigation scenarios across years 2016 - 2018 and find no qualitative difference in the result between years (see the supplementary materials).

**Figure 6.**
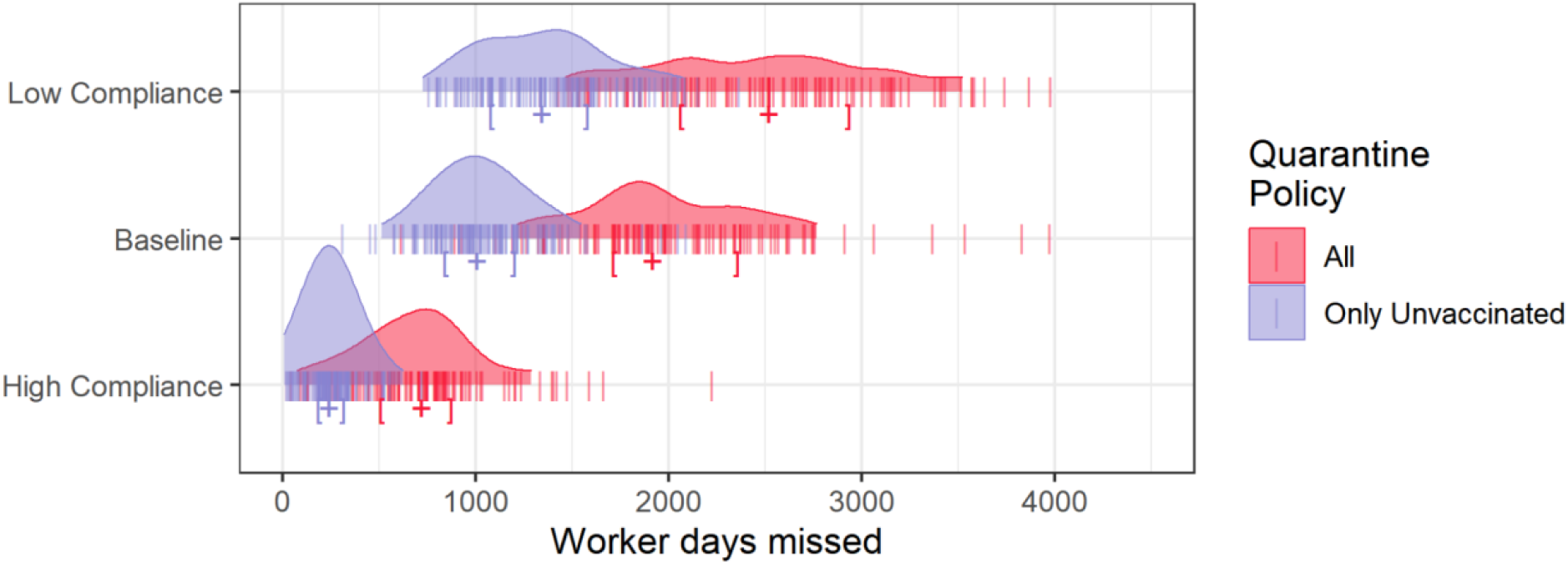
The distribution of worker days missed by scenario. The red denotes all workdays missed by vaccinated and unvaccinated firefighters while the blue denotes workdays missed by only unvaccinated firefighters. The Only Unvaccinated indication captures current guidance. Brackets indicate the interquartile range and plus signs indicate the median value for each distribution.

## Discussion

Our results suggest that vaccination and disease spread mitigations reduce the total number of SARS-CoV-2 infections in the wildland fire community, as well as reducing the number of infectious assignments and infectious reassignments to wildland fires. In addition, vaccination and disease spread mitigations lower the probability of outbreaks on individual fires and reduce workforce absenteeism. In our results we observe many more infections incurred off-fire than while firefighters are on assignment and similarly, more infectious assignments than reassignments. We do observe differential risk levels for crew personnel and management personnel. Below we discuss the implications of these results on the wildland firefighting system, as well as discussing some of the mechanisms that may be driving these results.

The national wildland firefighting system relies on scalable mobilization of individuals and groups of individuals from around the nation, and these individuals may serve in different roles and capacities depending on their qualifications and the needs of the incident. The population structure at a fire incident and its evolution over time as resources are mobilized/demobilized creates complex networks of interaction such that every incident carries different degrees of transmission risk. Fire personnel can be mobilized from all around the country, including reassignments from other incidents, such that there are systemic interdependencies in risk of transmission and potential for cascading outbreaks. In summary, the structure and function of the wildland firefighting system pose a unique set of risks from COVID-19, requiring a tailored approach to characterizing those risks.

Our primary focus here was analyzing potential COVID-19 impacts to workforce health and capacity, a topic of growing importance as increasing fire activity is expected to further strain the response capabilities of the system^17^. There are three primary workforce-related factors to consider. First, not captured in our analysis but worth mentioning, management of COVID-19 creates additional workload burden including screening/testing, isolating/quarantining, and interfacing with entities such as local public health agencies and hospitals – and this burden increases with the number of infections and outbreaks. Second, worker absenteeism due to isolation/quarantine requires greater coordination and prioritization of scarce resources both within and across incidents, and in some cases results in unfilled resource requests and understaffed incidents^10^. Depending on the degree of scarcity and substitutability of the affected resources^18^, this could result in inability to implement preferred strategies and tactics (e.g., lack of crews) or incident management organizations operating outside of their typical span of control (e.g., lack of key management personnel). Third, and perhaps most important to the workforce, missed days can translate into loss of assignments and loss of pay. For some of the firefighting workforce, the bulk of their annual salary comes from their time on assignment when their pay rate is increased due to overtime hours and hazard pay. In some cases, due to minimum personnel requirements for certain assignments, entire crews could be deemed unqualified if only some members of their team are in isolation or quarantine. Vaccination in such cases would insure against crew members having to quarantine due to exposure and would make more crews generally more available for assignments.

This point naturally leads to the primary finding of this analysis, that high vaccination rates in combination with the policy that vaccinated individuals do not need to quarantine after exposure results in significantly fewer worker days missed compared to other scenarios. The best case (High Compliance scenario with current quarantine policy) results in on average more than five times fewer missed worker days than the worst case (Low Compliance scenario without quarantine policy). Hence the importance of capturing uncertainty around vaccination uptake in the risk assessment and more broadly the importance of vaccination in maintaining system capacity.

Further, model results suggest that vaccination and disease spread mitigations reduce both infections and workforce absenteeism in the wildland fire community. There are two primary mechanisms at play: 1) vaccination and spread mitigation efforts keep infections low, leading to fewer isolations and 2) vaccinations allow exposed personnel to avoid quarantine. The contact structure of our ABM accounted for organizational structure and social distancing mitigations, and the ABM also captured heterogeneity in quarantine requirements according to individual agent and module status. The contact structure also led to the finding that infection risks may be higher for personnel that cannot “module as one.”

ABM results also show that most infections incurred by wildland firefighting personnel are likely to be from off fire sources rather than being incurred while on assignment. This implies that vaccination and mitigation techniques may prevent large outbreaks that cascade across the fire system, even in most Low Compliance scenarios. In other words, although the normal functioning of the system creates a systemic risk through reliance on a highly transient workforce with complex and dynamic exposure patterns, vaccination and social distancing on-fire can disrupt cascading outbreaks and effectively mitigate those systemic risks.

In addition to the ABM being useful for examining the spread of SARS-CoV-2 within and across fire incidents, it can also be used to simulate the spread of other respiratory diseases. It is documented that spread of “camp crud” (a generic term for any respiratory disease that spreads between personnel on wildland fire assignments) occurs on an annual basis. The results from this research have implications for the spread of a variety of infectious diseases, and the impact of the COVID-19 mitigation measures used herein may decrease disease and absenteeism from a variety of respiratory pathogens including influenza and RSV^19–21^. There could also be similarities with other dynamic populations such as emergency response or disaster relief where the ABM could prove useful. In addition, this ABM might also be repurposed for a variety of other applications in fire, ranging from optimal coordination and routing of aircraft to individual crew member movement and engagement in containment activities.

As with any modeling study, results are influenced by simplifications, assumptions, and parameter setting. We documented all model choices and the code is available for readers interested in exploring alternative parameters or different behavioral scenarios. The pandemic and its response to it will continue to be a fluid environment, for instance emerging variants or recent policy changes in the USA regarding mask wearing. This analysis does not capture all possible futures nor is it intended to be in any way considered as predictive.

What is intended is that results can be used to gain insight into how SARS-CoV-2 could spread in the wildland firefighting community, and how effective vaccination and social distancing may be at protecting workforce health and preventing workforce capacity degradation. Continuing to encourage personnel to mitigate the spread of SARS-CoV-2 through upcoming fire seasons is still important, even as vaccination rates rise. In addition, any method that raises vaccination rates may be an effective way to limit lost worker days both by mitigating disease spread and by preventing exposed individuals from having to isolate.

## Methods

In this section we describe our agent-based model in detail, describe the development of scenarios, and define the metrics we use to evaluate the effects of fire seasons and the spread of SARS-CoV-2 on the wildland fire workforce.

### Agent-based simulation model

Assignments to fires and spread of infection between personnel at those fires is simulated using an epidemiological ABM, with the viral states and transmission probabilities tailored to reflect SARS-CoV-2 spread through a wildland fire incident. In this model, each firefighter is modeled as an individual agent. Each individual is assigned a viral state each day. There are five viral states to which an individual may be assigned: susceptible, exposed, infectious-symptomatic, infectious-asymptomatic, and recovered. Susceptible individuals are assumed to have no immunity to the virus gained by previous SARS-CoV-2 exposure. Exposed individuals are those that have been exposed to the virus at a high enough viral load that they will become infectious. Infectious individuals are classified as symptomatic or asymptomatic. Symptomatic individuals have (or will eventually) develop COVID-19 symptoms and are able to spread the virus to others. Asymptomatic individuals are infectious but never show symptoms; these individuals are capable of spreading SARS-CoV-2 to others, but due to a lower viral load have a lower rate of transmission than those who are symptomatic^22^. Recovered individuals are those who have recovered from infection and are no longer able to spread SARS-CoV-2 to others.

The model simulates transmission of SARS-CoV-2 between firefighters (the agents in the model) through contact with other firefighters for each ongoing wildland fire; on-fire exposure occurs when susceptible individuals come into contact with an infectious individual. Transmission between fires occurs when exposed or infectious individuals leave one fire and are subsequently reassigned to another fire within the infectious period. When individuals are not on a fire, their probability of becoming infected is driven by prevalence-dependent geographic area specific parameters. While an individual will not spread infection to any other firefighters during the period they are off assignment, they can catch SARS-CoV-2 off-fire. They may then arrive at their next assignment exposed or infectious, at which point they may become a source of infection for that fire. Once an individual is exposed the model simulates the infected individual’s progression through the viral states. See left panel in Figure 2 for an illustration of the possible paths individuals may take through viral states.

In addition to tracking the daily viral state of each individual, the model also tracks each individual’s vaccination state. Our model has two vaccination states: vaccinated and unvaccinated. At the beginning of each simulation run a pre-specified number of firefighters is designated as vaccinated (dependent on the scenario). Over the course of the simulation an additional pre-specified number of individuals are vaccinated; personnel in isolation cannot become vaccinated, but individuals can get vaccinated while in any viral state. To model vaccine efficacy, a pre-specified proportion of susceptible individuals move from their current viral state directly to recovered, reflecting that those individuals cannot become infected or spread infection to others. The vaccinated individuals who do not move into the recovered class are still considered susceptible and are able to acquire and spread SARS-CoV-2.

Prior to 2020 the assumption of homogenous mixing among wildland fire personnel was appropriate, as personnel typically ate and slept in a confined area (typically called “fire camp”) as well as interacting at the location of planning and logistical activities (typically called “incident command post”). The fire camp and incident command post have provided conditions where other infectious diseases have spread with ease^3,4^. However, in response to the COVID-19 pandemic, several mitigation measures were developed to minimize contact between wildland fire personnel at large fire incidents. One of these measures was referred to as “module-as-one”^5^. This is a specific form of social distancing, by which crews seek to minimize all contact outside of their own crew. This practice is expected to continue through the 2021 fire season. To simulate module-as-one behavior, we grouped personnel associated with a specific crew or piece of ground equipment into a single module. Because management personnel are not able to be part of a module due to their duties requiring them to interact with a higher number of people, though at lower contact intensities, we treated them differently than crew personnel. These management personnel are the fire managers, planners and logistics personnel who spend most of their time at the incident management post. Thus, all personnel assigned to a management role are considered to be a single module in our model, and that module has different spread parameters associated with it. This logic for organizing personnel results in many 4-20 person modules on each fire in addition to a single management module which reflects the actual module structure on wildland fire incidents in 2020. In our simulation, one set of individuals is designated as the module leaders, and those are the only individuals that have contacts with others outside of their module (see right panel in Figure 2 for an example). Specifically, the leaders of each module contact only the leaders of the other modules. We assume four leaders per module for all crew modules. The management module has substantially more leaders than other modules because management personnel are regularly interacting with module leaders. Leadership status is randomly assigned to the individuals within a module each time a module mobilizes to an incident and stays constant for the duration of the assignment. Generally, modules move throughout the fire season intact, with few changes to personnel within the module. However, individuals on modules can change. Any individuals mobilized to a fire who are not assigned to an engine or crew module are classified as management; this is consistent with actual fire ordering practice. Contact between module members is substantially higher than contact between leaders, as module members are assumed to eat, sleep, and socialize together. Therefore, we provide a parameter to increase transmission between module members; we assume transmission between module members (both crew and management) is four times higher than transmission between leaders.

To facilitate our analyses of workforce impacts, we model and track which individuals are in isolation or quarantine each day. When a symptomatic individual recognizes that they have been infected, they isolate themselves from all other firefighters. The diagnosis of symptoms is not assumed to be immediate upon entry to the infectious state: our model includes a parameter to specify the average amount of time it takes for personnel to recognize their symptoms and a parameter to specify the average percentage of individuals who ever correctly diagnose their symptoms. For those individuals who do enter isolation, we base our isolation procedures on CDC guidelines^23^: once an individual is identified as infectious they isolate for 10 days or until 24 hours after symptoms are gone (i.e., one day after they move to the recovered state), whichever is longer. To reflect current practice, if an individual on a module isolates, all other individuals in their module must quarantine, regardless of viral state. Our model assumes full isolation during quarantine (i.e., isolated individuals do not transmit disease to any other individuals). The exception to this is the management modules; for those modules only the diagnosed individual is isolated. While in reality asymptomatic individuals could be isolated in response to a positive test, we did not include testing of non-symptomatic individuals in our simulations as that is not expected to be routine for firefighting personnel on all assignments. The model does isolate vaccinated persons, to allow us to account for the effect of vaccination on numbers of isolated personnel.

Because this quarantine policy is fairly strict, spread of SARS-CoV-2 from one fire to another can only occur under a very specific set of circumstances. First, the infected individual must be reassigned during their infectious window and must subsequently expose another individual at their new assignment prior to any isolation or quarantine. For symptomatic individuals, this is less likely to occur than for asymptomatic individuals, as a percentage of symptomatic individuals are assumed to recognize their symptoms and move into isolation. This percentage of symptomatic individuals who move into isolation is dependent on the level of mitigation compliance. For asymptomatic individuals, this fire to fire spread is more likely to occur as the only reason they would move into quarantine is if another module member is symptomatic and has recognized their symptoms. For viruses with a short period of time of symptomless infectiousness, few asymptomatic cases, and easily diagnosable symptoms, this isolation and quarantine policy would make reassignments that spread disease quite unlikely. However, COVID-19 has been associated with infectiousness prior to symptom onset^24^, a substantial proportion of asymptomatic cases^25^, and symptoms that may be attributable to the smoky conditions and physical exertion that firefighters regularly encounter^8^, thus, even with a strict isolation and quarantine policy SARS-CoV-2 may spread from one fire to another.

Our simulations use personnel assignment data from three historical fire seasons (2016-2018) to represent a range of possible outcomes for the coming fire season. Each individual simulation covers a single year and provides a possible disease spread outcome for that fire season. On the first day of the season (the day of the first assignment in our data) the probability of each individual being in an initial viral state is driven by a set of predetermined parameters (see the supplementary materials for specifics). The model then steps through each day in the fire season, checking daily on each individual’s assignment, module, and role and simulating and tracking individuals’ daily viral, vaccination, and isolation/quarantine status. Individuals’ contacts with others in their module and leaders’ contact with each other is modeled on each incident; an average number of infection-spreading contacts is calculated for each module and the group of leaders. This average number of infectious contacts that lead to a new infection is used as the mean of a Poisson distribution that is used to randomly assign to each individual on that module the number of successful infectious contacts they had that day. Any individual assigned one or more infection-producing close contacts with an infectious individual becomes exposed. Individuals who are off fire may contact SARS-CoV-2 with a probability dependent upon local transmission. Exposed and infectious individuals’ states are re-evaluated daily, and individuals move from exposed to symptomatic or asymptomatic and from symptomatic or asymptomatic to recovered based upon the daily probability of changing viral states (see supplementary material for specifics). In addition to daily re-evaluation of infectious states, individuals are also assessed for isolation. Symptomatic individuals are assigned to be in isolation based upon a random draw. Individuals within the same module as an isolated individual are then quarantined. Individuals move out of isolation and vaccination occurs based upon the isolation and vaccination methods described above. A detailed description of the ABM algorithm, the distributions used for draws and the associated parameters can be found in the supplementary materials. We simulate the model 100 times in each scenario (Baseline, High Compliance, Low Compliance) for each fire season to illustrate the uncertainty due to stochastic transmission. The simulation model and supporting functions were developed using R^26^ are available as an R package^27^.

### Scenario development

We developed scenarios to address two key uncertainties in the interplay between the fire season and the COVID-19 pandemic: the spatio-temporal variation of fire occurrence and uncertainty around vaccination and social distancing behaviors of wildland fire personnel. We address the variation in fire occurrence patterns by using fire assignments from three distinct fire seasons: 2016, 2017, and 2018. These years cover a range of spatial and temporal demand for wildland fire suppression resources. We address the uncertainty around vaccination and social distancing using a set of three behavioral scenarios.

To build our fire assignment dataset, we identified the set of large wildland fires (i.e., fires assigned a Type 1, Type 2, Type 3, National, or Area Command incident management team or incident commander) that burned in the US in 2016, 2017, and 2018 using data archived in the Resource Ordering and Status System (see ^3,9,10,28^ for previous peer reviewed studies using this data). Using these data we can track individuals uniquely across the fire season, identifying their daily assignments to large fires, the role they play on those fires, and the geographic area within which they are working. The 2016 fire season was a fairly average season, with slightly fewer fires and slightly fewer acres burned than the ten-year average, though the number of structures burned was slightly over the yearly average (calculated since 1999; NICC 2016). The 2017 fire season was a more severe fire season than 2016, with the number of acres burned well above the ten-year average as well as setting the record for most structures burned (1999-2017; NICC 2017). The 2018 fire season was also an above average fire season, with the number of acres burned well above the ten-year average and again setting a record for the number of structures burned (1999-2018; NICC 2018). Our assignment data matches these trends; the data include 190, 233, and 234 large fires in 2016, 2017 and 2018, respectively, with 37,299, 43,360, and 40,593 personnel assigned to at least one of these fires in 2016, 2017, and 2018, respectively. Further exploration of the fire assignment data can be found in the supplementary materials.

We address the uncertainty in vaccination of wildland fire personnel and compliance to social distancing behaviors by creating the three distinct behavioral scenarios that are described in the Results section (Baseline, Low Compliance, and High Compliance). The specific parameters used for each scenario can be found in the supplementary materials.

The calibration of the parameters representing the reproductive capacity of the virus are presented in detail in the supplementary materials. We aimed to have a median reproductive number for SARS-CoV-2 of 1.8, 1.34 and 0.8 people infected by a single infectious person for the Low Compliance, Baseline and High Compliance scenarios, respectively. These reproductive numbers assume an R_0_ of 2.4^29^, with the low compliance, baseline, and high compliance scenarios representing, respectively, a 25%, 44% and 67% reduction in transmission compared to uncontrolled transmission.

### Workforce impact evaluation metrics

Cumulative infections were counted daily for each simulation run. Infections are attributed by assignment status (i.e., off fire or on fire) and personnel role (crew or management) to allow for in-depth exploration of infection patterns. Infections can also be attributed to specific fires; for each run we counted the number of cases of SARS-CoV-2 on each fire. If a fire saw at least two cases from different crew modules, two management personnel with cases, or a combination of crew and management personnel with cases, we counted that fire as having an outbreak for that run. Cases did not have to be incurred on the fire, but the infected personnel had to be assigned to the fire, infectious, and not quarantined for at least one day.

The systematic risks of disease transmission to and across fires is assessed by counting the number of infectious assignments attributed to personnel who caught the disease off-fire as well as the number of infectious reassignments from personnel who leave one fire infectious and are subsequently reassigned to another fire while still infectious. Because infections can be attributed to personnel role, we can examine differential risks that occur across roles in addition to comparing risks of infection source.

When a symptomatic member of a firefighter module is diagnosed with COVID-19, all unvaccinated personnel within that module must quarantine, which implies few cases of COVID-19 may result in a substantial loss of workforce capacity. Therefore, a key metric we report is the number of cumulative days that firefighters were assigned to fires but were in isolation and thus limited in their ability to work; we refer to this as “worker days missed.” Only those who are unvaccinated would be asked to isolate, but we also track the number of individuals who would have been asked to isolate if they were not vaccinated to show the effect vaccination may have on workforce capacity. Therefore, we report two sets of worker days missed: 1) all isolated individuals (including those who are vaccinated) and 2) only isolated individuals who are not vaccinated. Because we track individuals in our model, a single day of isolation for a crew of 20 people results in 20 worker days missed.

## Conclusions

The COVID-19 pandemic poses a unique set of risks to the nationally interconnected wildland firefighting system. Infections on one fire have the potential to spread to other fires. Uncertainty around the upcoming fire season and behavioral choices regarding vaccination and mitigation of infection spread compounded the problem of understanding how COVID-19 might affect the wildland firefighting system in 2021 and beyond. Our model shows that current mitigation efforts limit the risk of transmission within a fire and across fires over the season. High behavioral compliance (i.e., high uptake of vaccines and adherence to spread mitigations) result in substantially fewer infections and lost worker days. These results reinforce the importance of continuing spread mitigations into future fire seasons and emphasize the value of vaccination within the workforce.

## Data Availability

The datasets of annual wildland firefighting assignments that were generated for this study are available from the corresponding author upon reasonable request; we chose not to make these publicly available due to the potential for the identification of individuals based upon their assignment history. The simulation model and supporting functions are available as an R package.

## Data Availability

The datasets of annual wildland firefighting assignments that were generated for this study are available from the corresponding author upon reasonable request; we chose not to make these publicly available due to the potential for the identification of individuals based upon their assignment history. The simulation model and supporting functions are available as an R package^27^.

## Additional Information

The author(s) declare no competing interests.

The findings and conclusions in this paper are those of the author(s) and should not be construed to represent any official USDA or U.S. Government determination or policy. USDA authors do not convey any copyright on this publication.

This manuscript is currently undergoing peer review and has not yet been fully vetted by referees external to the USDA.

## Acknowledgements

This research was supported by the U.S. Department of Agriculture (USDA) Forest Service and was funded in part by joint venture agreement number 18-JV-11221636-099 between Colorado State University and the USDA Forest Service Rocky Mountain Research Station. Additional funding was provided by the Joint Fire Science Program, which provided funds to the USDA Forest Service Rocky Mountain Research Station (agreement 20-S-01-2) to support this work at Colorado State University through the existing joint venture agreement.

## Author contributions

EJB, MPT, JB and AB wrote the wrote the main manuscript text. EJB, MPT, JB, AB, and JD developed the agent-based model. JD coded the model. EJB collected the data. JD, EJB, and JB prepared the figures. All authors reviewed the manuscript.

## Technical Appendix

Here in the technical appendix we include a detailed outline of the agent-based algorithm, a list of the specific parameters we used within the algorithm with sources for the parameters cited (where applicable), a description of how we processed the assignment data and a summary of the 2016-2018 fire seasons including comparisons between model runs, sample size comparisons, and a description of our calibration of the reproductive parameters we used for SARS-CoV-2.

### Agent-based model algorithm

1. Assign agents to their current incident and module.
  a. Agents not on an assignment are assigned to off-fire status.
  b. Leadership status is assigned for agents in modules on new assignments.
2. Simulate contacts between agents in each module, including the management modules. For each module:
  a. Identify all agents who are not isolated and are infectious (symptomatic or asymptomatic).
  b. Identify all agents who are not isolated and are susceptible or exposed.
  c. Given at least one infectious, not isolated agent in the module and at least one susceptible agent, calculate average number of infectious contacts (*a*_*m,d*_) that occur for an individual in module *m* on day *d* using the following equation, where *β*_*sym*_ is the daily transmission number for symptomatic agents, *β*_*asym*_ is the daily transmission number for asymptomatic agents, *c* is the contact intensity multiplier for modules, and *n*_*s,m,d*_ is the number of agents in the module *m* in state *s* on day *d*.

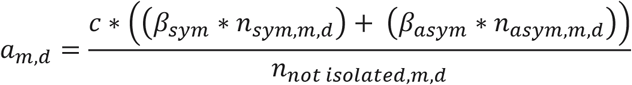
  d. For each susceptible agent, simulate the number of infectious contacts from other module members today using a draw from a Poisson distribution with the mean of the distribution (lambda) equal to *a*_*m,d*_. The viral state is changed from susceptible to exposed for each agent whose random draw is greater than zero.
3. Model contacts between module leaders
  a. Identify all module leaders who are not isolated and are infectious (symptomatic or asymptomatic)
  b. Identify all module leaders who are not isolated and are susceptible or exposed
  c. Given at least one infectious, not isolated leader and at least one susceptible leader, calculate calculate average number of infectious contacts (*a*_*l,d*_) that occur for an individual designated as a leader (*l*) on day *d* using the following equation where *β*_*sym*_ is the daily transmission number for symptomatic agents, *β*_*asym*_ is the daily transmission number for asymptomatic agents, and *n*_*s,l,d*_ is the number of agents designated as leaders in state *s* on day *d*.

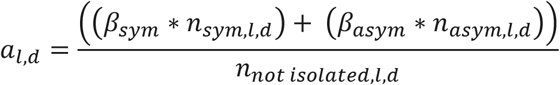
  d. For each susceptible leader, simulate the number of infectious contacts from other leaders today using a draw from a Poisson distribution with the mean of the distribution (lambda) equal to *a*_*l,d*_. The viral state is changed from susceptible to exposed for each leader whose random draw is greater than zero.
4. Model infectious contacts for off-fire agents.
  a. Identify all off-fire agents.
  b. For each agent, draw a random number from a Uniform(0,1) distribution.
  c. Compare the random draws to the off-fire infection parameter (*eir*). If the random draw is less than the off-fire infection parameter and the agent is susceptible, then the agent’s viral state changes to exposed.
5. Some exposed agents become infectious.
  a. Identify all agents in the exposed viral state.
  b. For each agent, draw two random numbers from a Uniform(0,1) distribution. The first draw will be used to determine if the agent is leaving the exposed state. The second draw will be used to determine which infectious state (symptomatic or asymptomatic) the agent enters, if they do leave the exposed state.
  c. Compare the random draw values to the daily probability of leaving the exposed state (*p*_*e*_=1/*D*_*e*_), where *D*_*e*_ is the average incubation period, and the probability of being symptomatic (*p*_*i*_). There are three possible outcomes for the agent:
    i. If the first random draw is less than the daily probability of leaving the exposed state and the second random draw is less than the probability of being symptomatic, then the agent’s viral state changes to symptomatic.
    ii. If the first random draw is less than the daily probability of leaving the exposed state and the second random draw is greater than or equal to the probability of being symptomatic, then the agent’s viral state changes to asymptomatic.
    iii. If the first random draw is greater than or equal to the daily probability of leaving the exposed state, then the agent’s viral state does not change.
6. Some infectious agents recover
  a. Identify all agents in the symptomatic or asymptomatic viral state.
  b. For each agent, draw a random number from a Uniform(0,1) distribution.
  c. Compare the random draw to the daily probability of recovering (*p*_*r*_). If the random draw is less than the daily probability of recovery, then the agent’s viral state changes to recovered.
7. Some infectious agents isolate.
  a. Identify all symptomatic and asymptomatic agents who are not currently isolated.
  b. For each agent, draw a random number from a Uniform(0,1) distribution.
  c. Compare the random draw value to the daily probability that an agent correctly identifies their symptoms and/or receives a positive test (*p*_*IQ*_ for symptomatic agents and *p*_*AQ*_ for asymptomatic agents). If the random draw is less than the daily probability that an agent correctly identifies their symptoms and/or receives a positive test then the agent’s isolation state changes to isolated.
8. Isolate other agents, increment isolation day counts, and release agents from isolation.
  a. Identify all isolated agents.
  b. If the agent is on an assignment and part of a crew module, then isolate all other non-isolated module members.
  c. For all isolated agents, increment the number of days they have been isolated by one.
  d. If an agent has been isolated for greater than the number of required isolation days (*D*_*q*_) and their viral state is not symptomatic then their isolation state changes to not isolated and their isolation day count is set to 0. If the agent is still symptomatic after the required number of isolation days, then they continue to be isolated until they move to the recovered state (driven by *p*_*r*_, the daily probability of recovery).
9. Vaccinate agents.
  a. Identify all agents that are not isolated and not vaccinated.
  b. Randomly sample these agents to determine who is vaccinated on that day, vaccinating exactly the number of agents specified for that geographic area on that day (*vo*_*g,d*_ for management and *vno*_*g,d*_ for crew).
  c. Sample the newly vaccinated agents whose viral state is susceptible to determine if their viral state changes from susceptible to recovered, changing the states for exactly the number specified by the vaccination efficacy parameter (*ve* * *vo*_*g,d*_ and *ve* * *vno*_*g,d*_).
10. If this is not the last day of the season, increment forward one day and go back to step 1. If this is the final day of the season, save the following information for each resource on each day of the season: viral state, vaccination state, isolation state, and leader status.

### Parameters used in the agent-based model

The parameters used in the simulations are listed in Tables 1 and 2. Where appropriate, we also list references for parameter values.

**Table 1:**
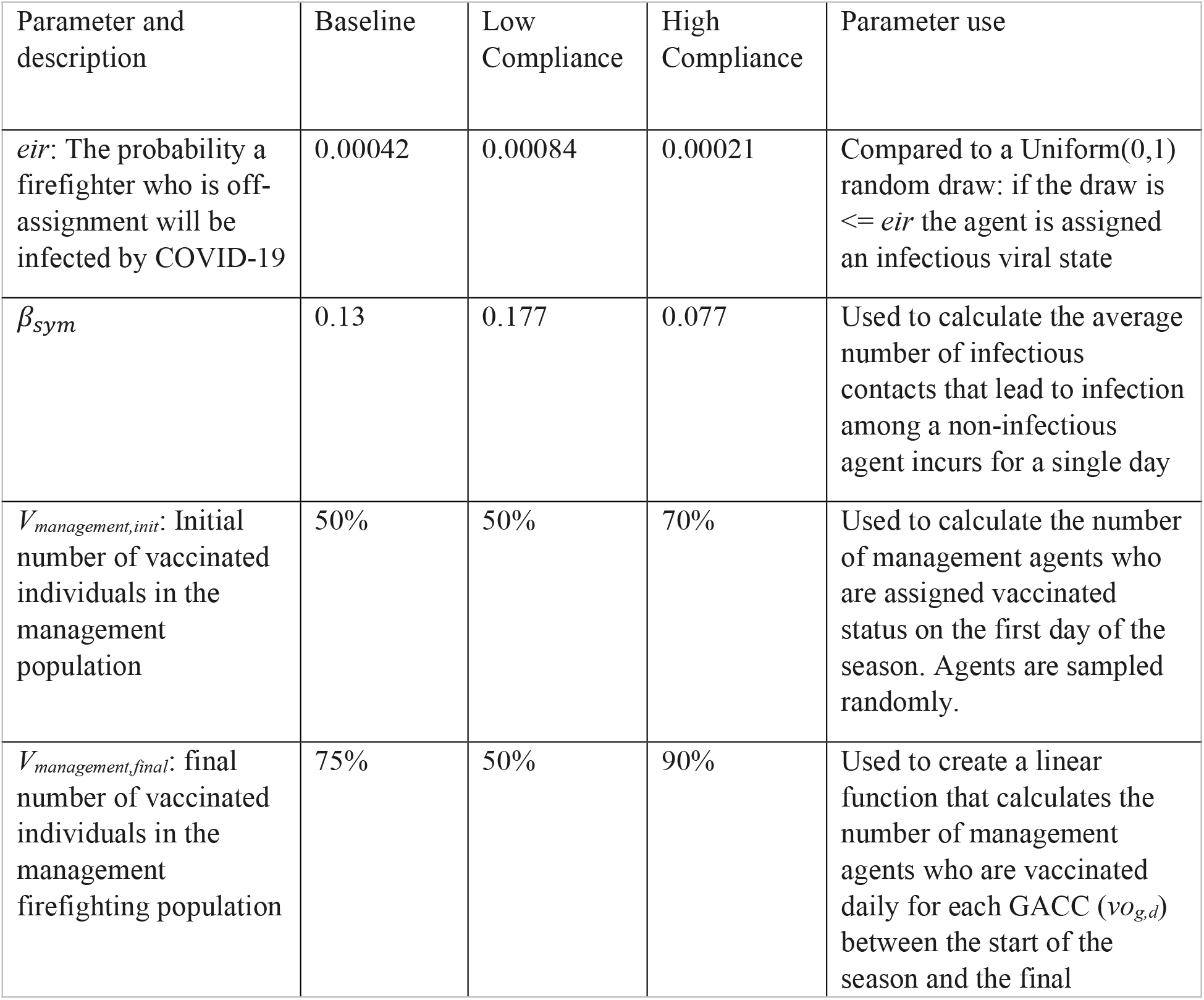
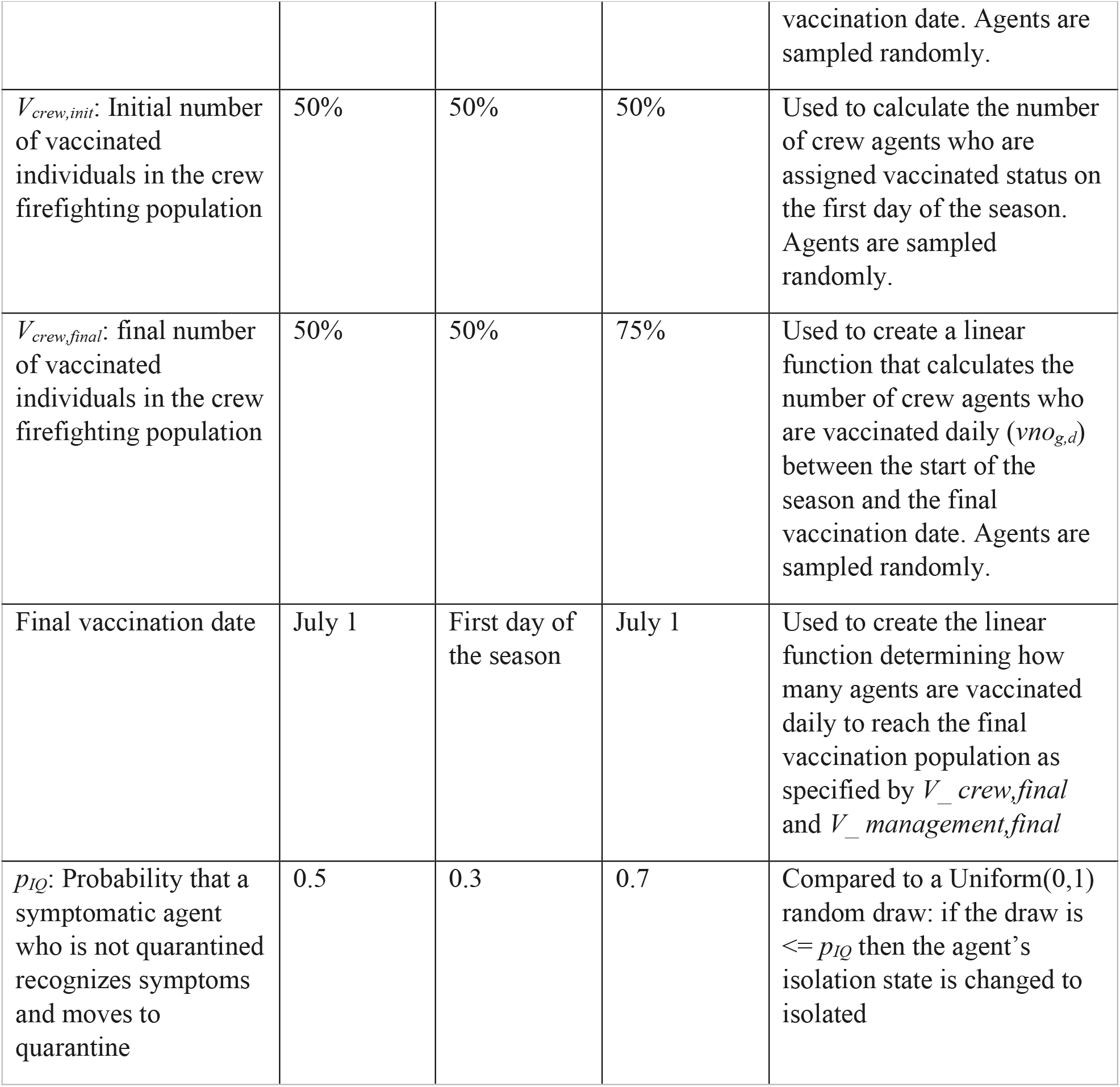
Parameter values that vary across scenarios

**Table 2:** Parameter values that stay constant across scenarios

| Parameter | Value | Description | Parameter use |
| --- | --- | --- | --- |
| $p_i$ | 0.4286 | Probability that an infectious agent is symptomatic (weighted by population age)<sup>1</sup> | Compared to a Uniform(0,1) random draw: if the draw is $\leq p_i$ then the agent's viral state is changed to symptomatic, else the viral state is changed to asymptomatic |
| $p_{AQ}$ | 0 | Proportion asymptomatic that quarantines due to a positive test | Compared to a Uniform(0,1) random draw: if the draw is $< p_{AQ}$ then the agent's isolation state is changed to isolated |
| $R_{init}$ | 25% | Initial percent of recovered individuals in the firefighting population | Used to calculate the number of agents who start the season in the recovered viral state. Agents are sampled randomly. |
| $Num\ leaders$ | 4 | Number of leaders per module (crew) | Calculates the number of agents who are assigned leadership status for crew modules. |
| $I_{init}$ | 0.598% | Initial percent of infectious individuals in the firefighting population | Used to calculate the number of agents who start the season in the symptomatic viral state. Agents are sampled randomly. |
| $D_e$ | 5 | Average incubation period<sup>2,3</sup> | Used to calculate $p_e$, the probability of leaving the exposed state, which is compared to a Uniform(0,1) random draw: if the draw is $\leq p_e$ then the agent's viral state will change to either symptomatic or asymptomatic |
| $D_r$ | 8 | Average days from onset of infectiousness to recovery<sup>4,5</sup> | Used to calculate $p_r$, the probability of leaving an infectious state, which is compared to a Uniform(0,1) random draw: if the draw is $\leq p_r$ then the agent's viral state will change to recovered |
| $l$ | 0.5 | Proportion management designated leaders | Calculates the number of agents who are assigned leadership status for management modules. |
| $c$ | 4 | Reproduction multiplier for modules | Used to calculate the average number of infectious contacts that a non-infectious agent incurs for a single day ($a_{m,d}$) |
| $D_q$ | 10 | Required days of isolation<sup>6</sup> | Compared to days an agent has been in isolation: given days in isolation is $\geq D_q$ and agent is recovered, agent leaves isolation state. |
| $ve$ | 0.95 | Vaccine efficacy<sup>7,8</sup> | Used to calculate the number of agents who move from symptomatic to recovered upon receiving their vaccine. |

### Assignment data and fire season summaries

To build our fire assignment dataset, we identified the set of wildland fires that were managed by a Type 1, Type 2, Type 3, National, or Area Command incident management team or incident commander which burned in the US in 2016, 2017, and 2018 using data archived in the Resource Ordering and Status System (see Thompson et al 2020, Belval et al 2020, 2016, Lyon et al 2018 for previous peer reviewed studies using this data). For each of these fires we obtained the assignments for each of the individuals that provided wildland firefighting capacity or wildland fire management capacity on the fire. Individuals are assigned unique identifiers that are constant across the season, allowing us to identify which days each individual was assigned to each of the large fires and to observe movement of individuals between large fires across the fire season. The home geographic area for each individual is typically provided; if the home geographic area is unknown then personnel are assigned to the geographic area in which they are first assigned to an incident. Individuals are also classified by their role on the fire, that is, they are assigned to a specific hand crew, a crew managing a piece of equipment (for example, engine or dozer), or management (personnel handling the logistics and planning for the fire). We use these roles to create the modules on each fire. We did not include personnel assigned to aerial resources as aircraft assignments in ROSS are not always reliable (Belval et al 2020) and those personnel have a lower level of contact with other firefighting personnel.

The 2016 fire season was a fairly average season, with slightly fewer fires and slightly fewer acres burned than the ten-year average, though the number of structures burned was slightly over the yearly average (calculated since 1999). There was a pulse of fire activity early in the season (pre-July) driven by the SW and Southern California, followed by a pause (early July), followed by a medium level of fire activity throughout the rest of the season. The 2016 fire season was unique in having a pulse of destructive fire activity in the Southern Area in December, which accounted for over 2000 of the 4312 structures burned in 2016 (NICC 2016). The 2017 fire season was a more severe fire season than 2016, with the number of acres burned well above the ten-year average as well as setting the record for most structures burned (1999-2017). The Northern Rockies, Great Basin, and Northwest all saw substantial fire activity mid-season (August-September). Southern California experienced a very destructive pulse of fire in December (NICC 2017). The 2018 fire season was also an above average fire season, with the number of acres burned well above the ten-year average and again setting a record for the number of structures burned (1999-2018). The Northwest, Great Basin, and California experienced substantial fire activity mid-season (July-August) and California again experienced a late season pulse of fire in November (NICC2018). The number of personnel on assignment daily from each Geographic Area is shown in Figure S.1. The assignment data include 190, 233, and 234 large fires in 2016, 2017 and 2018, respectively, with 37,299, 43,360, and 40,593 personnel assigned to at least one of these fires in 2016, 2017, and 2018, respectively.

**Figure S.1:**
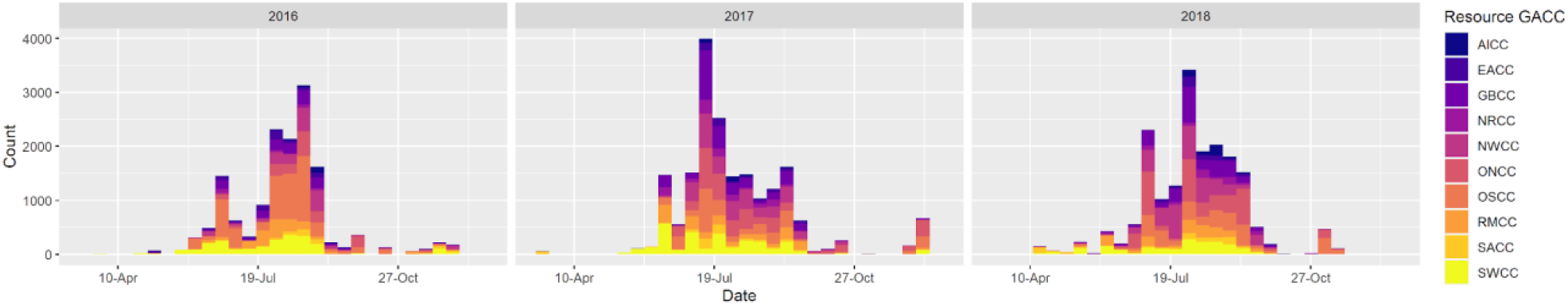
Number of potential disease spreading reassignments across the 2016, 2017, and 2018 fire season. The color indicates the geographic area from which the firefighter originates. Abbreviations for geographic areas (GACCs) are: AICC (Alaska), EACC (Eastern Area), GBCC (Great Basin), NRCC (Northern Rockies), NWCC (Northwest), ONCC (Northern California), OSCC (Southern California), RMCC (Rocky Mountain Area), SACC (Southern Area), SWCC (Southwest).

Simulations using the Baseline scenario parameters show similar distributions of both infection prevalence over time, cumulative infections (Figure S.1a), and worker days missed (Figure S.1b). The median number of cumulative infections for runs using the 2016, 2017, and 2018 assignments under the Baseline scenario assumptions was 79.5 [IQR: 72-88], 94 [IQR: 81-102], and 94 [IQR: 82.75-108.25] respectively. We do observe a higher level of cumulative infections overall in 2017 and 2018 than 2016; this is because the total number of personnel assigned to a large fire is highest in 2017, leading to a larger pool of personnel that can be infected off fire. The median number of cumulative infections in 2016 was 1498 [IQR: 1471-1521], in 2017 was 1915 [IQR: 1892-1944], and in 2018 was 1808 [IQR: 1782-1849]. Similar to the number of on-fire infections, the number of worker days missed (both when quarantining all personnel and when quarantining only vaccinated individuals) does not vary substantially between years; worker days missed (for quarantining only unvaccinated individuals) was 995.1 [IQR: 801-1244.5], 1007 [IQR: 842-1198] and 1003.5 [IQR: 810.5-1193.5] for 2016, 2017 and 2018, respectively.

**Figure S.2:**
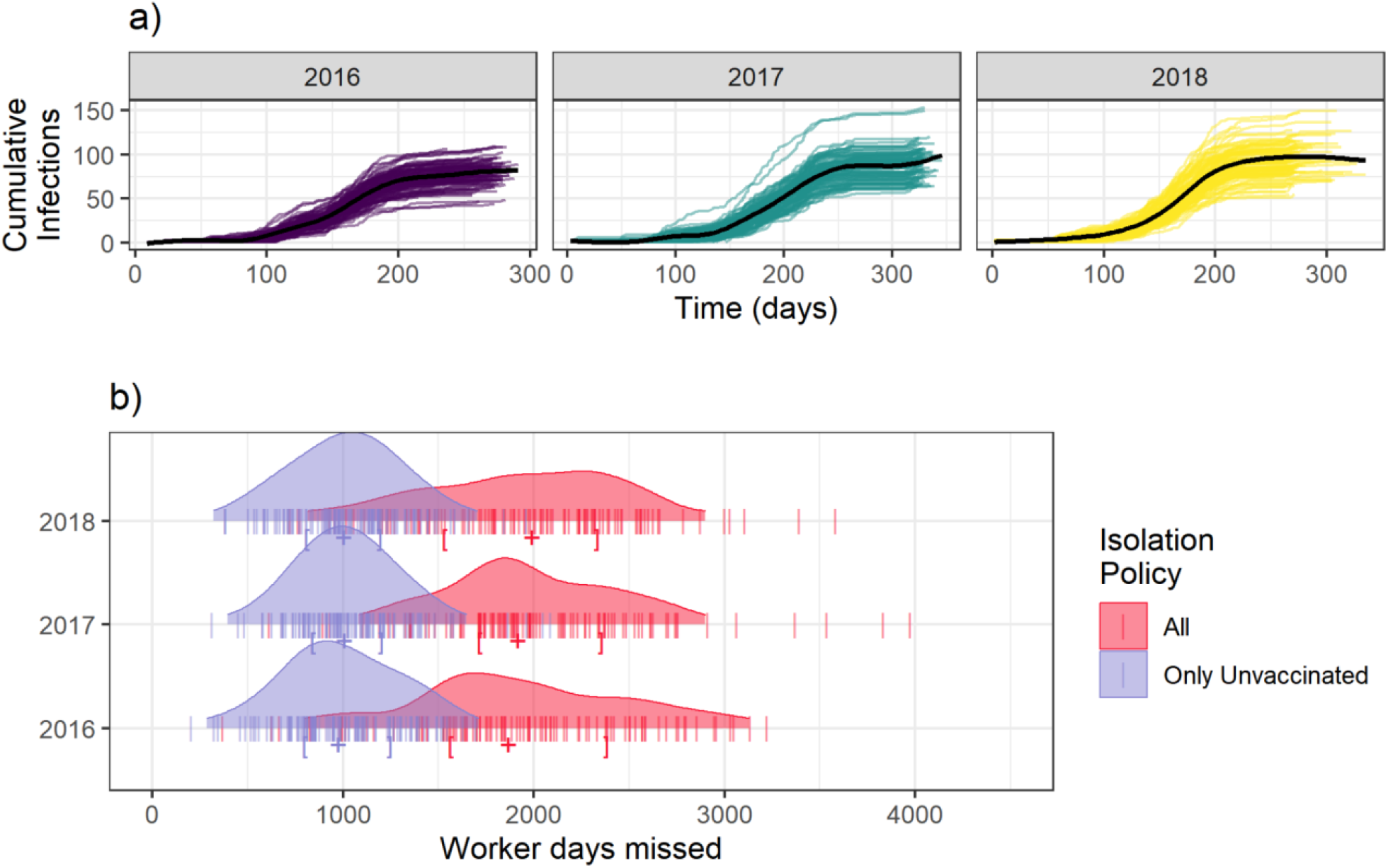
Prevalence paths for cases of SARS-CoV-2 incurred during a fire assignment (a) and the distribution of worker days missed for (b) using the Baseline scenario parameters occurring for runs using 2016, 2017 and 2018 assignment data.

### Calibration of reproductive parameters for SARS-CoV-2

A key calculation in the model is the average number of infectious individuals that a single individual contacts at a close enough level to transmit the virus. This can be calculated using the equation from step 1c (reproduced below for ease of reading), where *a*_*m,d*_ is the average number of infectious contacts that occur for an individual in module *m* on day *d, n*_*sym,m,d*_ and *n*_*asym,m,d*_ are the number of symptomatic personnel and number of asymptomatic personnel, respectively, in module *m* on day *d* who are not isolated and *n*_*not isolated,m,d*_ is the total number of not isolated personnel in module *m* on day *d*. This includes a “reproduction multiplier” (*c*) that allows the intensity of contact to vary depending upon whether the contact is between module members or leaders: contact is substantially higher within modules.

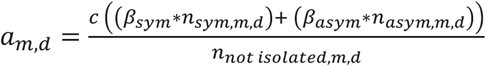

Based on estimates of disease spread from those who are asymptomatic relative to those who are symptomatic, we assumed 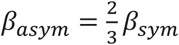. To calibrate *β*_*sym*_ and *β*_*asym*_we ran the model on the five days with the maximum number of personnel from three fires from 2017 that have previously been used to examine COVID-19 spread through wildland fire personnel: the Lolo Peak fire, the Highline fire, and the Tank Hollow fire. For the Low Compliance scenario, we aimed to provide a reproductive capacity such that each single infectious individuals would infect 1.8 other individuals on average. For the High Compliance scenario, we aimed to provide a reproductive capacity such that each single infectious individual would infect 0.8 other individuals on average. For the Baseline scenario, we aimed to provide a reproductive capacity such that each single infectious individual would infect 1.34 other individuals on average. The final values of *β*_*sym*_ that we used are listed in Table 2.

### Sample size for Monte Carlo simulations

We wanted to run enough simulations to adequately represent the stochasticity within this system, but to also use computing resources efficiently. We ran sample sets of simulations using both 100 runs and 500 runs. We found the distributions of infections did not vary substantially between 100 and 500 runs (see Table S.3 for comparisons).

**Table S.3:** A comparison of the distribution of cumulative infections from a set of 500 simulation runs and a set of 100 simulation runs using the 2017 assignment data for all three behavioral scenarios.

| Scenario | Cumulative infections 500 runs | Cumulative infections 100 runs |
| --- | --- | --- |
| Low Compliance | 4506 [IQR: 4454-4556] | 4512 [IQR: 4461-4566] |
| Baseline | 1916 [IQR: 1885-1950] | 1915 [IQR: 1892-1944] |
| High Compliance | 635 [IQR: 619-653] | 634.5 [IQR: 618.8-654.2] |

## Notes

### Competing Interest Statement

The authors have declared no competing interest.

### Author Declarations

This research did not qualify as "research involving human subjects or clinical investigation involving human subjects" according to the Colorado State University IRB as we did not collect health data on individuals.

## References

1. Moore, P. et al. Wildland fire management under COVID-19. Brief 1, review of materials. Wageningen University, The Netherlands. (2020) https://doi.org/10.18174/521344.

2. Stoof, C. R., Poortvliet, M., Hannah, B., Steffens, R. & Moore, P. Preview Brief 2: Wildland Fire Management under COVID-19, Survey Results. Wageningen University, The Netherlands. (2020) https://doi.org/10.18174/522586.

3. Thompson, M. P., Bayham, J. & Belval, E. Potential COVID-19 Outbreak in Fire Camp: Modeling Scenarios and Interventions. Fire 3, 38 (2020).

4. National Wildfire Coordinating Group. Infectious Disease Guidance for Wildland Fire Incidents, Emergency Medical Committee. https://www.nwcg.gov/committees/emergency-medical-committee/infectious-disease-guidance (2020).

5. National Wildfire Coordinating Group. Guidance for Prevention and Management of COVID-19 During Wildland Fire. https://www.nwcg.gov/partners/fmb/guidance-prevention-management (2021).

6. Khubchandani, J. et al. COVID-19 Vaccination Hesitancy in the United States: A Rapid National Assessment. J Community Health 46, 270–277 (2021).

7. Navarro, K. M. et al. Wildland firefighter exposure to smoke and COVID-19: A new risk on the fire line. Science of The Total Environment 760, 144296 (2021).

8. Managing a COVID-19 Worst-Case Scenario The Cameron Peak Fire Story. https://experience.arcgis.com/experience/0d12d48a842745868c1cef3b7b99cd83/page/page_0/ (2021).

9. Belval, E. J. et al. Studying interregional wildland fire engine assignments for large fire suppression. Int. J. Wildland Fire 26, 642–653 (2017).

10. Belval, E. J., Stonesifer, C. S. & Calkin, D. E. Fire Suppression Resource Scarcity: Current Metrics and Future Performance Indicators. Forests 11, 217 (2020).

11. Wallentin, G., Kaziyeva, D. & Reibersdorfer-Adelsberger, E. COVID-19 Intervention Scenarios for a Long-term Disease Management. Int J Health Policy Manag 1 (2020) doi:10.34172/ijhpm.2020.130.

12. Firth, J. A., Klepac, P., Kissler, S., Kucharski, A. J. & Spurgin, L. G. Using a real-world network to model localized COVID-19 control strategies. Nat Med 26, 1616–1622 (2020).

13. Li, J. & Giabbanelli, P. J. Identifying Synergistic Interventions to Address COVID-19 Using a Large Scale Agent-Based Model. Preprint at http://medrxiv.org/lookup/doi/10.1101/2020.12.11.20247825 (2020) xdoi:10.1101/2020.12.11.20247825.

14. Rockett, R. J. et al. Revealing COVID-19 transmission in Australia by SARS-CoV-2 genome sequencing and agent-based modeling. Nat Med 26, 1398–1404 (2020).

15. Jalayer, M., Orsenigo, C. & Vercellis, C. CoV-ABM: A stochastic discrete-event agent-based framework to simulate spatiotemporal dynamics of COVID-19. 2007.13231 [physics] (2020).

16. Holmdahl, I., Kahn, R., Hay, J. A., Buckee, C. O. & Mina, M. J. Estimation of Transmission of COVID-19 in Simulated Nursing Homes With Frequent Testing and Immunity-Based Staffing. JAMA Netw Open 4, e2110071 (2021).

17. Abatzoglou, J. T., Juang, C. S., Williams, A. P., Kolden, C. A. & Westerling, A. L. Increasing Synchronous Fire Danger in Forests of the Western United States. Geophys Res Lett 48, (2021).

18. Stonesifer, C. S., Calkin, D. E. & Hand, M. S. Federal fire managers’ perceptions of the importance, scarcity and substitutability of suppression resources. Int. J. Wildland Fire 26, 598 (2017).

19. Olsen, S. J. et al. Decreased influenza activity during the COVID-19 pandemic—United States, Australia, Chile, and South Africa, 2020. Am J Transplant 20, 3681–3685 (2020).

20. Varela, F. H. et al. Absence of detection of RSV and influenza during the COVID-19 pandemic in a Brazilian cohort: Likely role of lower transmission in the community. J Glob Health 11, 05007 (2021).

21. Soo, R. J. J., Chiew, C. J., Ma, S., Pung, R. & Lee, V. Decreased Influenza Incidence under COVID-19 Control Measures, Singapore. Emerg. Infect. Dis. 26, 1933–1935 (2020).

22. Li, R. et al. Substantial undocumented infection facilitates the rapid dissemination of novel coronavirus (SARS-CoV-2). Science 368, 489–493 (2020).

23. Centers for Disease Control and Prevention. When You Can be Around Others After You Had or Likely Had COVID-19. https://www.cdc.gov/coronavirus/2019-ncov/if-you-are-sick/end-home-isolation.html (2021).

24. Sanche, S. et al. High Contagiousness and Rapid Spread of Severe Acute Respiratory Syndrome Coronavirus 2. Emerg. Infect. Dis. 26, 1470–1477 (2020).

25. Buitrago-Garcia, D. et al. Asymptomatic SARS-CoV-2 infections: a living systematic review and meta-analysis. Preprint at http://medrxiv.org/lookup/doi/10.1101/2020.04.25.20079103 (2020) xdoi:10.1101/2020.04.25.20079103.

26. R Core Team. R: A language and environment for statistical computing. R Foundation for Statistical Computing, Vienna, Austria (2019).

27. Dilliott, J. jakedilliott/covidfireMASS: First Official Release. Zenodo (2021). doi:10.5281/ZENODO.4990871.

28. Lyon, K. M., Huber-Stearns, H. R., Moseley, C., Bone, C. & Mosurinjohn, N. A. Sharing contracted resources for fire suppression: engine dispatch in the Northwestern United States. Int. J. Wildland Fire 26, 113 (2017).

29. Wu, J. T., Leung, K. & Leung, G. M. Nowcasting and forecasting the potential domestic and international spread of the 2019-nCoV outbreak originating in Wuhan, China: a modelling study. The Lancet 395, 689–697 (2020).

## References

1. Davies, N. G. et al. Age-dependent effects in the transmission and control of COVID-19 epidemics. Nat Med 26, 1205–1211 (2020).

2. Lauer, S. A. et al. The Incubation Period of Coronavirus Disease 2019 (COVID-19) From Publicly Reported Confirmed Cases: Estimation and Application. Annals of Internal Medicine 172, 577– 582 (2020).

3. Bi, Q. et al. Epidemiology and transmission of COVID-19 in 391 cases and 1286 of their close contacts in Shenzhen, China: a retrospective cohort study. The Lancet Infectious Diseases 20, 911–919 (2020).

4. He, X. et al. Temporal dynamics in viral shedding and transmissibility of COVID-19. Nat Med 26, 672–675 (2020).

5. Cheng, H.-Y. et al. Contact Tracing Assessment of COVID-19 Transmission Dynamics in Taiwan and Risk at Different Exposure Periods Before and After Symptom Onset. JAMA Intern Med 180, 1156 (2020).

6. Centers for Disease Control and Prevention. When You Can be Around Others After You Had or Likely Had COVID-19. https://www.cdc.gov/coronavirus/2019-ncov/if-you-are-sick/end-home-isolation.html (2021).

7. Baden, L. R. et al. Efficacy and Safety of the mRNA-1273 SARS-CoV-2 Vaccine. N Engl J Med 384, 403–416 (2021).

8. Polack, F. P. et al. Safety and Efficacy of the BNT162b2 mRNA Covid-19 Vaccine. N Engl J Med 383, 2603–2615 (2020).

